# Alzheimer’s Related Neurodegeneration Mediates Air Pollution Effects on Medial Temporal Lobe Atrophy

**DOI:** 10.1101/2023.11.29.23299144

**Authors:** Andrew J. Petkus, Lauren E. Salminen, Xinhui Wang, Ira Driscoll, Joshua Millstein, Daniel P. Beavers, Mark A. Espeland, Meredith N. Braskie, Paul M. Thompson, Ramon Casanova, Margaret Gatz, Helena C. Chui, Susan M Resnick, Joel D. Kaufman, Stephen R. Rapp, Sally Shumaker, Diana Younan, Jiu-Chiuan Chen

## Abstract

Exposure to ambient air pollution, especially particulate matter with aerodynamic diameter <2.5 μm (PM_2.5_) and nitrogen dioxide (NO_2_), are environmental risk factors for Alzheimer’s disease and related dementia. The medial temporal lobe (MTL) is an important brain region subserving episodic memory that atrophies with age, during the Alzheimer’s disease continuum, and is vulnerable to the effects of cerebrovascular disease. Despite the importance of air pollution it is unclear whether exposure leads to atrophy of the MTL and by what pathways. Here we conducted a longitudinal study examining associations between ambient air pollution exposure and MTL atrophy and whether putative air pollution exposure effects resembled Alzheimer’s disease-related neurodegeneration or cerebrovascular disease-related neurodegeneration.

Participants included older women (n = 627; aged 71-87) who underwent two structural brain MRI scans (MRI-1: 2005-6; MRI-2: 2009-10) as part of the Women’s Health Initiative Memory Study of Magnetic Resonance Imaging. Regionalized universal kriging was used to estimate annual concentrations of PM_2.5_ and NO_2_ at residential locations aggregated to 3-year averages prior to MRI-1. The outcome was 5-year standardized change in MTL volumes. Mediators included voxel-based MRI measures of the spatial pattern of neurodegeneration of Alzheimer’s disease (Alzheimer’s disease pattern similarity scores [AD-PS]) and whole-brain white matter small-vessel ischemic disease (WM-SVID) volume as a proxy of global cerebrovascular damage. Structural equation models were constructed to examine whether the associations between exposures with MTL atrophy were mediated by the initial level or concurrent change in AD-PS score or WM-SVID while adjusting for sociodemographic, lifestyle, clinical characteristics, and intracranial volume.

Living in locations with higher PM_2.5_ (per interquartile range [IQR]=3.17µg/m^3^) or NO_2_ (per IQR=6.63ppb) was associated with greater MTL atrophy (β_PM2.5_ = −0.29, 95% confidence interval [CI]=[−0.41,-0.18]; β_NO2_ =-0.12, 95%CI=[−0.23,-0.02]). Greater PM_2.5_ was associated with larger increases in AD-PS (β_PM2.5_ = 0.23, 95%CI=[0.12,0.33]) over time, which partially mediated associations with MTL atrophy (indirect effect= −0.10; 95%CI=[−0.15, −0.05]), explaining approximately 32% of the total effect. NO_2_ was positively associated with AD-PS at MRI-1 (β_NO2_=0.13, 95%CI=[0.03,0.24]), which partially mediated the association with MTL atrophy (indirect effect= −0.01, 95% CI=[−0.03,-0.001]). Global WM-SVID at MRI-1 or concurrent change were not significant mediators between exposures and MTL atrophy.

Findings support the mediating role of Alzheimer’s disease-related neurodegeneration contributing to MTL atrophy associated with late-life exposures to air pollutants. Alzheimer’s disease-related neurodegeneration only partially explained associations between exposure and MTL atrophy suggesting the role of multiple neuropathological processes underlying air pollution neurotoxicity on brain aging.

## Introduction

Cognitive impairment leading to Alzheimer’s disease and related dementias disproportionately impacts women,^1^ and is among the leading causes of disability and death worldwide.^2^ Clinical dementia stems from underlying neurologic disease which lead to neurodegeneration several years prior to cognitive and functional deficits.^3^ The medial temporal lobe (MTL), consisting of the hippocampus, parahippocampal gyrus, entorhinal cortex, and amygdala, is an important brain structure subserving episodic memory.^4^ After age 70, healthy older adults lose approximately 1.5-2% of their MTL volume annually on average.^5^ Individuals with Alzheimer’s disease, the most common dementia etiology, experience more than double the rate of MTL atrophy compared to healthy older adults.^5^ Although given less attention, atrophy of the MTL likely also occurs in vascular dementia and stroke which is the second most common cause of dementia.^6–8^ Various neurodegenerative disease processes differentially impact the various MTL subregions. For example, the entorhinal cortex (ERC) is the first MTL subregion impacted by Alzheimer’s disease.^9^ Volumetric changes to the ERC may also be pronounced, compared to other MTL structures in normal aging,^10^ are associated with longitudinal memory changes in cognitively normal individuals,^11^ and may also be an important biomarker associated with other dementia etiologies.^12^ Identifying environmental factors that contribute to MTL atrophy during the preclinical disease stage is important for dementia prevention.

Accumulating evidence has shown that late-life exposure to ambient air pollution is an environmental risk factor for dementia.^13,14^ Despite the importance of the MTL across the various dementia etiologies, there have been few studies examining associations between exposure to air pollution in later-life on atrophy of the MTL with most studies providing mixed findings.^15–21^ The pathways by which air pollution exposure may lead to the atrophy of the MTL in cognitively healthy individuals are also unclear. These pathways may include Alzheimer’s disease-related neurodegeneration, cerebrovascular disease, or other neuropathological factors. Studies with animals and humans report a link between air pollution exposure and Alzheimer’s disease neuropathology.^22^ Our previous epidemiological work also reports a link between air pollution and increased Alzheimer’s disease-related neurodegeneration over a five-year period.^23,24^ A larger body of studies has demonstrated the toxic effects of air pollution exposure on the vascular system including a link between air pollution and cerebrovascular disease.^25,26^ Air pollution exposure may also exert a neurotoxic effect on the MTL through other pathways as studies report links between air pollution exposure and other neuropathological factors implicated with MTL atrophy.^27,28^ Examining if putative associations between air pollution exposure and MTL atrophy resemble patterns of neurodegeneration consistent with Alzheimer’s disease or cerebrovascular disease will provide a better understanding of the neurotoxic effects of air pollution on the brain.

To address the knowledge gaps, the current study examined the longitudinal associations between ambient air pollution exposure and MTL atrophy in older women without dementia. We then examined whether putative air pollution exposure effects resembled AD-related neurodegeneration or cerebrovascular disease-related neurodegeneration.

## Materials and methods

### Participants and study design

We analyzed longitudinal data from 627 community-dwelling older women who participated in the MRI substudy of the larger Women’s Health Initiative (WHI) Memory Study (WHIMS; n=7,479, age 65-79), which was an ancillary study of the WHI clinical trial of hormone therapy for postmenopausal women. Between April 2005 and January 2006, 1405 WHIMS participants, across 14 geographically diverse WHI centers, underwent a structural MRI scan (MRI-1). Between 2009-2010, 720 women completed a follow-up structural MRI scan (MRI-2). Since this study focused on mechanisms of exposure-related MTL atrophy at the preclinical stage of Alzheimer’s disease and related dementias, we excluded 11 women with probable mild cognitive impairment (MCI) or probable dementia at MRI-1 as determined through neuropsychological evaluation and clinical adjudication. Women with missing data on the mediators (n=64), and covariates (n=19) were also excluded, rendering a final analytic sample of 627 cognitively unimpaired older women. See **Figure 1** for a timeline of study assessments (Panel A) and a flowchart of study participation (Panel B).

**Figure 1.**
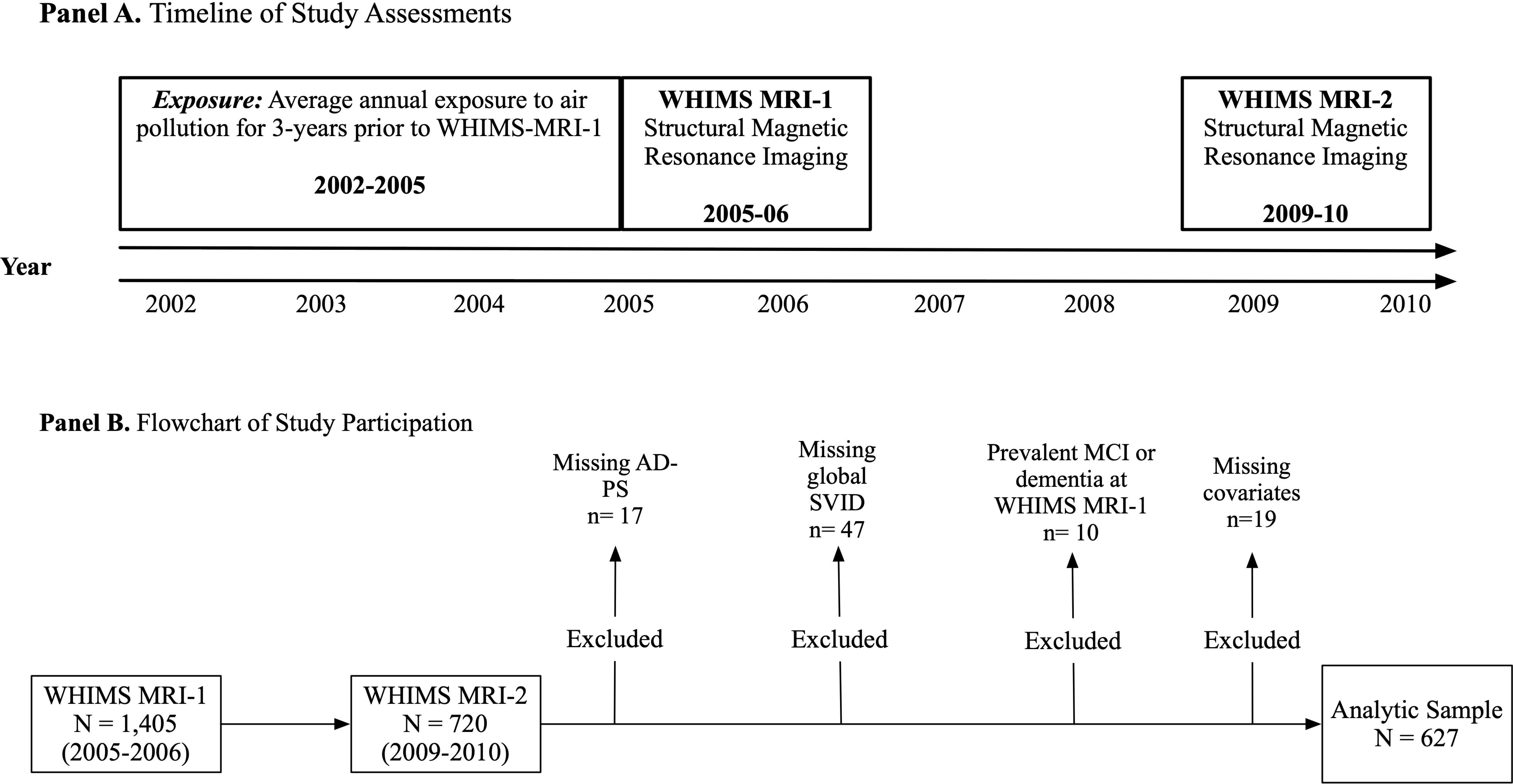
A timeline of study assessments is presented in Panel A while a flowchart of study participation is presented in Panel B.

### Air pollution exposure estimates

Participant residential address data were collected prospectively at each WHI assessment and geocoded.^29^ Regionalized universal kriging models^30,31^ were used to estimate the annual mean concentrations of ambient PM_2.5_ (in μg/m^3^) and NO_2_ (in ppb) at the participant’s residence for the 3 years prior to MRI-1. The models used US Environmental Protection Agency (EPA) monitoring data and over 300 geographic covariates in a partial least-squares regression to estimate air pollution exposure. The model includes geographic covariates (e.g., population density, distance to roads, and vegetation in the vicinity, etc.) to estimate exposures. These models are reliable and valid with high cross-validation with an R^2^ of 0.88 for PM_2.5_ and 0.85 for NO_2._ We calculated a 3-year weighted average of air pollution with the length of stay at each residential location within the 3-year time window as a weight to account for residential mobility.

### Structural MRI acquisition and processing

Participants completed two structural MRI brain scans on 1.5T scanners at one of 14 WHIMS clinical centers using a standardized protocol developed by the WHIMS-MRI Quality Control Center at the University of Pennsylvania. The scan series for volumetric imaging used a 22cm field of view and a 256×256 acquisition matrix and the following image series: oblique axial spin density/T2-weighted spin echo images, oblique axial fast fluid-attenuated inversion recovery (FLAIR) T2-weighted spin echo image and oblique axial fast spoiled 3D T1-weighted gradient echo images. Details about the pulse sequence parameters were published previously.^32,33^

MTL regions were extracted using a multi-atlas region segmentation (MUSE) that transformed site-specific labeled atlases into a harmonized map.^34,35^ MUSE follows a voxel-based spatial adaptation strategy to transform multiple atlases with different warping algorithms and regularization parameters into an ensemble-based parcellation of anatomical reference labels to harmonize the T1-weighted volumetric images. This approach leads to robust segmentation accuracy and is superior to other multi-atlas segmentation and label fusion methods, especially for multi-site MRI investigations and longitudinal analyses.^34,35^ Total MTL volume was operationally defined as the summed bilateral gray matter volumes of the hippocampus, amygdala, parahippocampal gyrus (PHG), and entorhinal cortex (ERC). We estimated total MTL atrophy and subregional MTL atrophy by calculating the volume changes from MRI-1 to MRI-2 and standardized this change by the number of years between the two visits. Numerical changes were then further standardized on a z-score metric based on the means and standard deviations of the sample.

### Assessment of Alzheimer’s disease-related neurodegeneration

We used Alzheimer’s disease pattern similarity (AD-PS) scores to estimate individual risk for Alzheimer’s disease-related neurodegeneration. AD-PS scores are metrics that reflect the degree of spatial similarity between a participant’s gray matter and the pattern of gray matter in individuals with Alzheimer’s disease. Scores were calculated using data-driven methods developed and validated using data from the Alzheimer’s disease neuroimaging initiative (ADNI).^36^ Briefly, a high-dimensional elastic net regularized logistic regression approach was used to derive a voxel-based model discriminating between cognitively unimpaired controls and those with clinically diagnosed dementia due to Alzheimer’s disease in ADNI based on gray matter probability maps. The class-conditional probabilities of membership to the Alzheimer’s disease group are called AD-PS scores. Discriminative maps derived from the model’s coefficients, which characterize the spatial pattern of gray matter differences, covered Alzheimer’s disease-vulnerable brain regions including the amygdala, hippocampus, PHG, thalamus, inferior temporal areas, and midbrain. Detailed technical information is provided elsewhere.^37^ The resulting AD-PS score can be interpreted as a measure of Alzheimer’s disease anatomical risk, with higher scores indicating higher Alzheimer’s disease risk. AD-PS scores were calculated at both MRI-1 and MRI-2. Our primary metrics of interest were AD-PS at MRI-1 and the change in AD-PS scores between the MRI visits.

### Assessment of Small Vessel Ischemic Disease

To assess the potential mediating role of white matter small vessel ischemic disease (WM-SVID), white matter lesions were segmented using a deep learning based segmentation, DeepMRSeg.^38^ White matter was classified as normal or “abnormal”, and abnormal voxels were summed across regions to represent a global measure of WM-SVID volume.

### Covariates of interest

At the WHI inception (1993–1998), a structured questionnaire was administered to gather information on the geographic region of residence (Northeast, South, Midwest, West), age, race/ethnicity (“White, non-Hispanic” and “Other ethnic backgrounds”), socioeconomic factors (education, family income, and employment status), and lifestyle factors (smoking status, alcohol use, and physical activity), and self-reported use of postmenopausal hormones. Body mass index (BMI) was determined from measured height and weight and depressive symptoms were recorded using the Center for Epidemiologic Studies Depression Scale-short form. Self-reported histories of cardiovascular disease (CVD; e.g., heart problems, problems with blood circulation, blood clots) and related risk factors (e.g., hypertension, treatment-dependent hypercholesterolemia, diabetes mellitus) were assessed using valid and reliable measures.^39,40^ As very few women endorsed a history of diabetes or hypercholesterolemia, we created a binary variable to indicate any versus no history of CVD to boost statistical power. In addition to the covariates collected at WHI inception, lifestyle factors, BMI, hypertension, and history of CVD were also updated at MRI-1. Socioeconomic status (SES) of residential neighborhoods was characterized using US Census tract-level data and estimated at both WHI inception and MRI-1^41^ ; a higher neighborhood SES score indicated a more socioeconomically-favorable neighborhood.

### Statistical analysis

Multivariable linear regressions were used to estimate the associations between air pollution exposures and MTL volume changes in total MTL volume while adjusting for sociodemographic variables, lifestyle factors, clinical characteristics, and intracranial volume. We estimated the association between PM_2.5_ and NO_2_ on MTL volume change in separate models. Structural equation models (SEMs) were used to test four putative mediators of exposure-related MTL volume change in separate models: (1) AD-PS score at MRI-1, (2) 5-year change in AD-PS, (3) SVID volume at MRI-1, and (4) 5-year change in SVID volume. The SEM mediation used standard methods to estimate direct and indirect exposure effects on MTL volumes over time (**Figure 2**). The direct effect was defined as the effect of exposures on MTL change independent of the hypothesized mediator (**Figure 2, path c**). The indirect effect was defined as the effect of exposures on MTL change explained by the hypothesized mediator, which is a product of the exposure effect on the mediator (**Figure 2, path a**) and the effect of the mediator on MTL changes (**Figure 2, path b**). The total effect of exposures on MTL volume changes is the sum of the direct and indirect effects. All estimated parameters were adjusted for sociodemographic, lifestyle, clinical covariates, and ICV (**Figure 2**). The 95% confidence interval (CI) of the indirect effect was calculated using 5,000 bootstrapped samples. Mediation analyses were conducted using the MPLUS Automation package^42^ in R.

**Figure 2.**
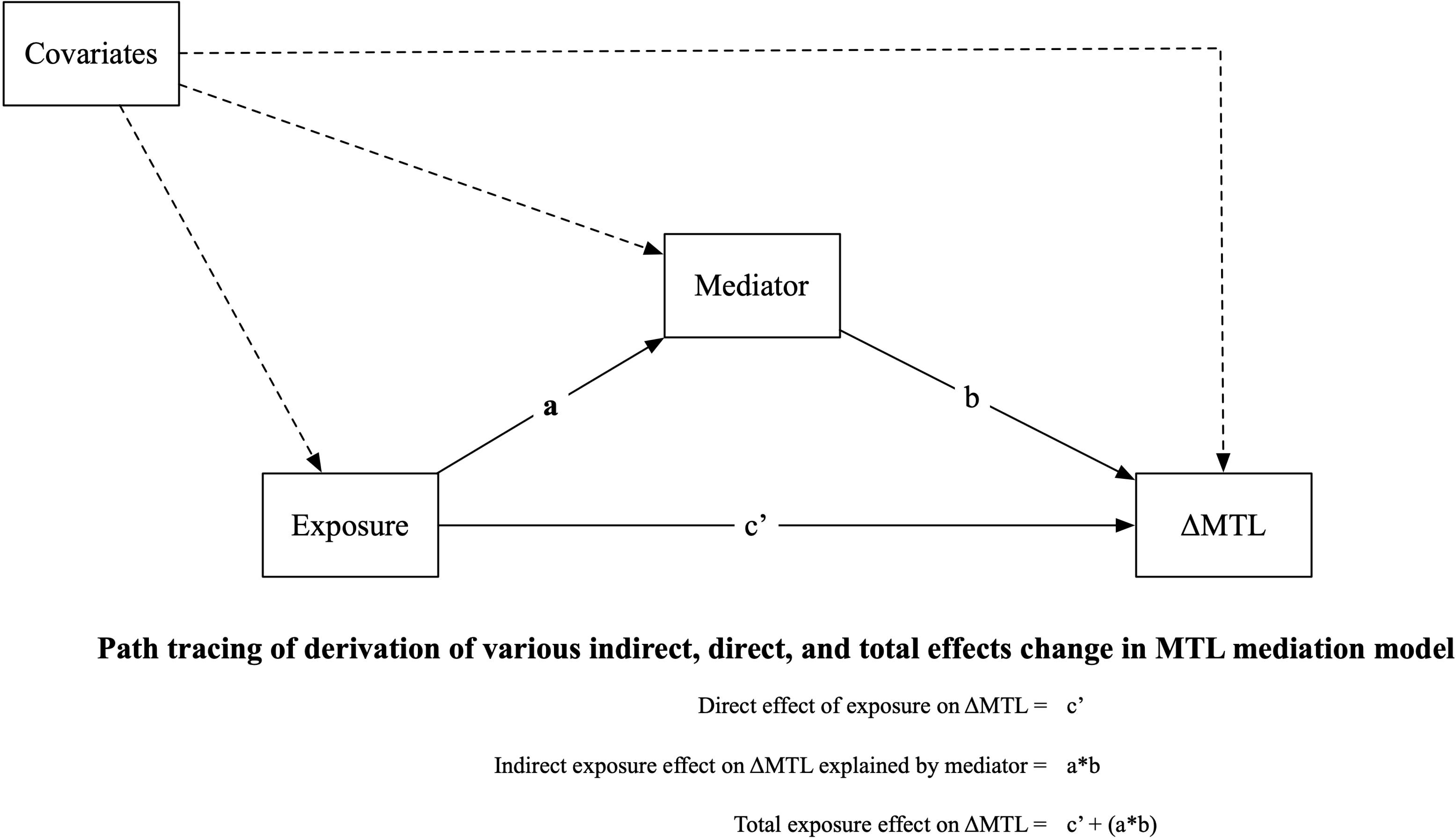
Simplified illustration of the structural equation model (SEM) to estimate whether Alzheimer’s disease neurodegeneration or global small-vessel-ischemic disease mediate observed associations between air pollution and medical temporal lobe (MTL) atrophy.

Exploratory analyses assessed whether the magnitude of the observed mediations differed across MTL subregions. We conducted these analyses because prior studies suggest that air pollution exposure may differentially impact the various MTL subregions.^21^ Additionally, studies suggest heterogeneous patterns of atrophy across the various MTL subregions across various neurodegenerative disease processes.

### Data availability

Access to all data elements used in this study may be made available following the established WHI policies.

## Results

### Descriptive characteristics of the sample

Participants were mostly non-Hispanic White women (*n=*592, 94%) with an average age of 77.42 ± 3.49 years. Table 1 compares the distribution of the 3-year average PM_2.5_ and NO_2_ exposures prior to MRI-1 by population characteristics. Compared to non-Hispanic White women, women who self-identified with other race/ethnic backgrounds tended to reside in locations with higher NO_2_ and PM_2.5_ exposures. Compared to women living in the South, women living in the Northeast and Midwest had higher PM_2.5_ exposures. Older women living in the South had lower NO_2_ exposures compared to those in other regions. Women who lived in the most socioeconomically-unfavorable neighborhoods at WHI inception or reported a history of postmenopausal hormone therapy had higher PM_2.5_ exposures compared to their counterparts, whereas women living in the most socioeconomically-favorable neighborhoods at WHI inception had higher NO_2_ exposures. Compared to their counterparts, higher NO_2_ exposures were also observed in current smokers, women who consumed one or more alcoholic drinks per day, or those with BMI<25 kg/m^2^.

**Table 1.**
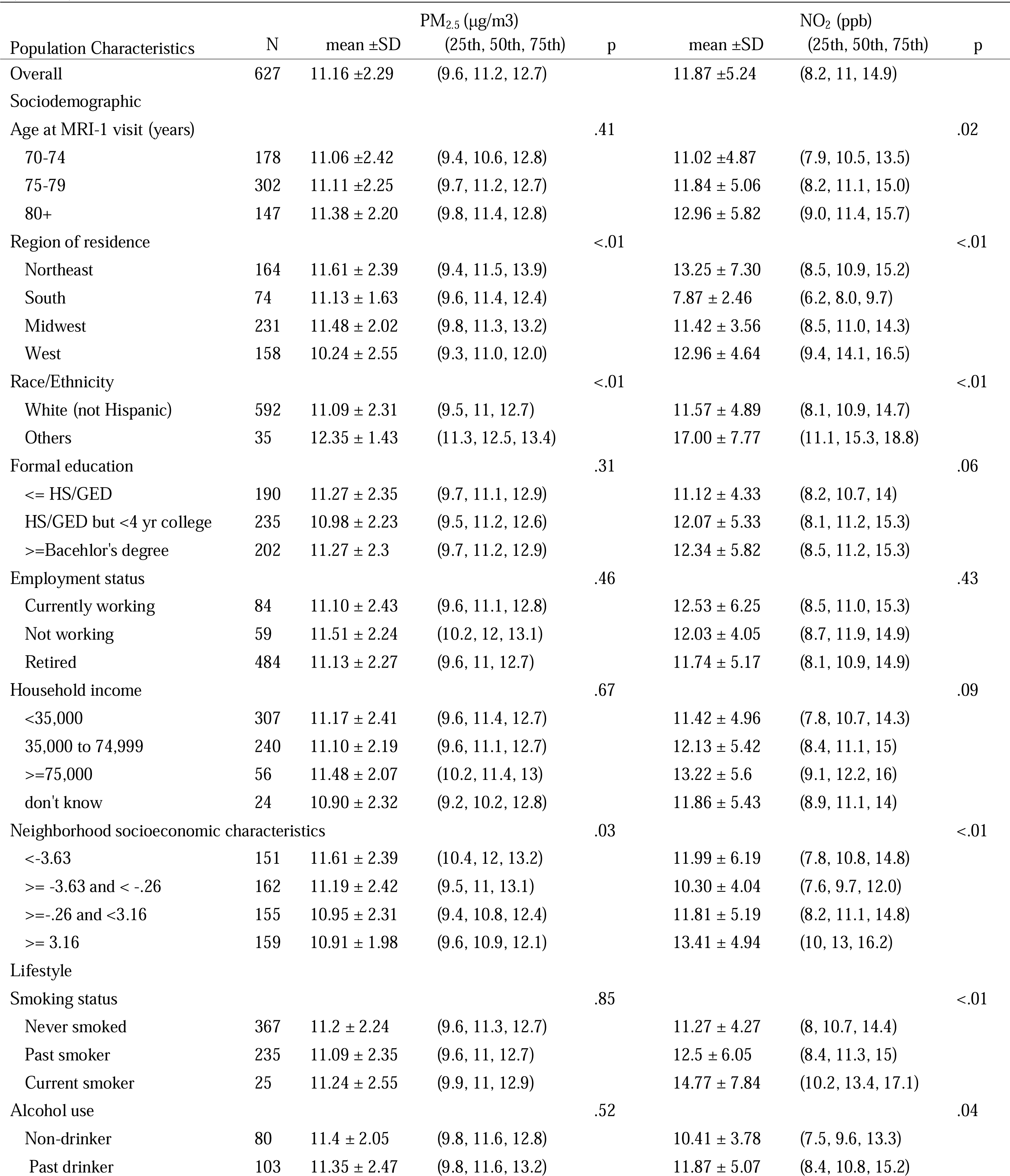
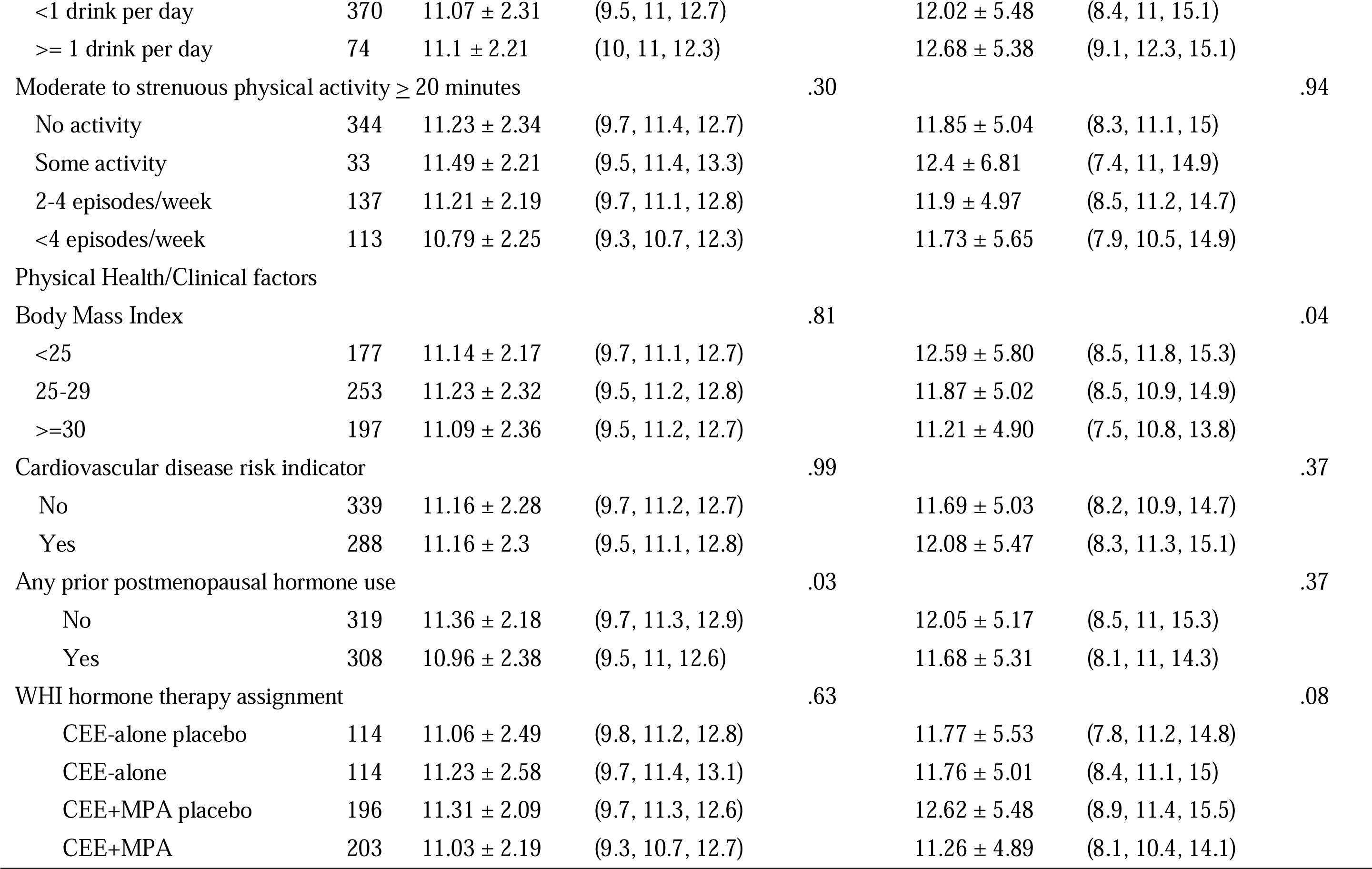
Distribution of air pollution exposures by population characteristics in the Women’s Health Initiative Memory Study MRI cohort (N =627)

| Population Characteristics | N | mean ±SD | PM<sub>2.5</sub> (µg/m3)<br>(25th, 50th, 75th) | p | mean ±SD | NO<sub>2</sub> (ppb)<br>(25th, 50th, 75th) | p |
| --- | --- | --- | --- | --- | --- | --- | --- |
| Overall | 627 | 11.16 ±2.29 | (9.6, 11.2, 12.7) |  | 11.87 ±5.24 | (8.2, 11, 14.9) |  |
| Sociodemographic |  |  |  |  |  |  |  |
| Age at MRI-1 visit (years) |  |  |  | .41 |  |  | .02 |
| 70-74 | 178 | 11.06 ±2.42 | (9.4, 10.6, 12.8) |  | 11.02 ±4.87 | (7.9, 10.5, 13.5) |  |
| 75-79 | 302 | 11.11 ±2.25 | (9.7, 11.2, 12.7) |  | 11.84 ± 5.06 | (8.2, 11.1, 15.0) |  |
| 80+ | 147 | 11.38 ± 2.20 | (9.8, 11.4, 12.8) |  | 12.96 ± 5.82 | (9.0, 11.4, 15.7) |  |
| Region of residence |  |  |  | <.01 |  |  | <.01 |
| Northeast | 164 | 11.61 ± 2.39 | (9.4, 11.5, 13.9) |  | 13.25 ± 7.30 | (8.5, 10.9, 15.2) |  |
| South | 74 | 11.13 ± 1.63 | (9.6, 11.4, 12.4) |  | 7.87 ± 2.46 | (6.2, 8.0, 9.7) |  |
| Midwest | 231 | 11.48 ± 2.02 | (9.8, 11.3, 13.2) |  | 11.42 ± 3.56 | (8.5, 11.0, 14.3) |  |
| West | 158 | 10.24 ± 2.55 | (9.3, 11.0, 12.0) |  | 12.96 ± 4.64 | (9.4, 14.1, 16.5) |  |
| Race/Ethnicity |  |  |  | <.01 |  |  | <.01 |
| White (not Hispanic) | 592 | 11.09 ± 2.31 | (9.5, 11, 12.7) |  | 11.57 ± 4.89 | (8.1, 10.9, 14.7) |  |
| Others | 35 | 12.35 ± 1.43 | (11.3, 12.5, 13.4) |  | 17.00 ± 7.77 | (11.1, 15.3, 18.8) |  |
| Formal education |  |  |  | .31 |  |  | .06 |
| <= HS/GED | 190 | 11.27 ± 2.35 | (9.7, 11.1, 12.9) |  | 11.12 ± 4.33 | (8.2, 10.7, 14) |  |
| HS/GED but <4 yr college | 235 | 10.98 ± 2.23 | (9.5, 11.2, 12.6) |  | 12.07 ± 5.33 | (8.1, 11.2, 15.3) |  |
| >=Bacehlor's degree | 202 | 11.27 ± 2.3 | (9.7, 11.2, 12.9) |  | 12.34 ± 5.82 | (8.5, 11.2, 15.3) |  |
| Employment status |  |  |  | .46 |  |  | .43 |
| Currently working | 84 | 11.10 ± 2.43 | (9.6, 11.1, 12.8) |  | 12.53 ± 6.25 | (8.5, 11.0, 15.3) |  |
| Not working | 59 | 11.51 ± 2.24 | (10.2, 12, 13.1) |  | 12.03 ± 4.05 | (8.7, 11.9, 14.9) |  |
| Retired | 484 | 11.13 ± 2.27 | (9.6, 11, 12.7) |  | 11.74 ± 5.17 | (8.1, 10.9, 14.9) |  |
| Household income |  |  |  | .67 |  |  | .09 |
| <35,000 | 307 | 11.17 ± 2.41 | (9.6, 11.4, 12.7) |  | 11.42 ± 4.96 | (7.8, 10.7, 14.3) |  |
| 35,000 to 74,999 | 240 | 11.10 ± 2.19 | (9.6, 11.1, 12.7) |  | 12.13 ± 5.42 | (8.4, 11.1, 15) |  |
| >=75,000 | 56 | 11.48 ± 2.07 | (10.2, 11.4, 13) |  | 13.22 ± 5.6 | (9.1, 12.2, 16) |  |
| don't know | 24 | 10.90 ± 2.32 | (9.2, 10.2, 12.8) |  | 11.86 ± 5.43 | (8.9, 11.1, 14) |  |
| Neighborhood socioeconomic characteristics |  |  |  | .03 |  |  | <.01 |
| <-3.63 | 151 | 11.61 ± 2.39 | (10.4, 12, 13.2) |  | 11.99 ± 6.19 | (7.8, 10.8, 14.8) |  |
| >= -3.63 and < -.26 | 162 | 11.19 ± 2.42 | (9.5, 11, 13.1) |  | 10.30 ± 4.04 | (7.6, 9.7, 12.0) |  |
| >=-.26 and <3.16 | 155 | 10.95 ± 2.31 | (9.4, 10.8, 12.4) |  | 11.81 ± 5.19 | (8.2, 11.1, 14.8) |  |
| >= 3.16 | 159 | 10.91 ± 1.98 | (9.6, 10.9, 12.1) |  | 13.41 ± 4.94 | (10, 13, 16.2) |  |
| Lifestyle |  |  |  |  |  |  |  |
| Smoking status |  |  |  | .85 |  |  | <.01 |
| Never smoked | 367 | 11.2 ± 2.24 | (9.6, 11.3, 12.7) |  | 11.27 ± 4.27 | (8, 10.7, 14.4) |  |
| Past smoker | 235 | 11.09 ± 2.35 | (9.6, 11, 12.7) |  | 12.5 ± 6.05 | (8.4, 11.3, 15) |  |
| Current smoker | 25 | 11.24 ± 2.55 | (9.9, 11, 12.9) |  | 14.77 ± 7.84 | (10.2, 13.4, 17.1) |  |
| Alcohol use |  |  |  | .52 |  |  | .04 |
| Non-drinker | 80 | 11.4 ± 2.05 | (9.8, 11.6, 12.8) |  | 10.41 ± 3.78 | (7.5, 9.6, 13.3) |  |
| Past drinker | 103 | 11.35 ± 2.47 | (9.8, 11.6, 13.2) |  | 11.87 ± 5.07 | (8.4, 10.8, 15.2) |  |
| <1 drink per day | 370 | 11.07 ± 2.31 | (9.5, 11, 12.7) |  | 12.02 ± 5.48 | (8.4, 11, 15.1) |  |
| ≥ 1 drink per day | 74 | 11.1 ± 2.21 | (10, 11, 12.3) |  | 12.68 ± 5.38 | (9.1, 12.3, 15.1) |  |
| Moderate to strenuous physical activity ≥ 20 minutes |  |  |  | .30 |  |  | .94 |
| No activity | 344 | 11.23 ± 2.34 | (9.7, 11.4, 12.7) |  | 11.85 ± 5.04 | (8.3, 11.1, 15) |  |
| Some activity | 33 | 11.49 ± 2.21 | (9.5, 11.4, 13.3) |  | 12.4 ± 6.81 | (7.4, 11, 14.9) |  |
| 2-4 episodes/week | 137 | 11.21 ± 2.19 | (9.7, 11.1, 12.8) |  | 11.9 ± 4.97 | (8.5, 11.2, 14.7) |  |
| <4 episodes/week | 113 | 10.79 ± 2.25 | (9.3, 10.7, 12.3) |  | 11.73 ± 5.65 | (7.9, 10.5, 14.9) |  |
| Physical Health/Clinical factors |  |  |  |  |  |  |  |
| Body Mass Index |  |  |  | .81 |  |  | .04 |
| <25 | 177 | 11.14 ± 2.17 | (9.7, 11.1, 12.7) |  | 12.59 ± 5.80 | (8.5, 11.8, 15.3) |  |
| 25-29 | 253 | 11.23 ± 2.32 | (9.5, 11.2, 12.8) |  | 11.87 ± 5.02 | (8.5, 10.9, 14.9) |  |
| ≥30 | 197 | 11.09 ± 2.36 | (9.5, 11.2, 12.7) |  | 11.21 ± 4.90 | (7.5, 10.8, 13.8) |  |
| Cardiovascular disease risk indicator |  |  |  | .99 |  |  | .37 |
| No | 339 | 11.16 ± 2.28 | (9.7, 11.2, 12.7) |  | 11.69 ± 5.03 | (8.2, 10.9, 14.7) |  |
| Yes | 288 | 11.16 ± 2.3 | (9.5, 11.1, 12.8) |  | 12.08 ± 5.47 | (8.3, 11.3, 15.1) |  |
| Any prior postmenopausal hormone use |  |  |  | .03 |  |  | .37 |
| No | 319 | 11.36 ± 2.18 | (9.7, 11.3, 12.9) |  | 12.05 ± 5.17 | (8.5, 11, 15.3) |  |
| Yes | 308 | 10.96 ± 2.38 | (9.5, 11, 12.6) |  | 11.68 ± 5.31 | (8.1, 11, 14.3) |  |
| WHI hormone therapy assignment |  |  |  | .63 |  |  | .08 |
| CEE-alone placebo | 114 | 11.06 ± 2.49 | (9.8, 11.2, 12.8) |  | 11.77 ± 5.53 | (7.8, 11.2, 14.8) |  |
| CEE-alone | 114 | 11.23 ± 2.58 | (9.7, 11.4, 13.1) |  | 11.76 ± 5.01 | (8.4, 11.1, 15) |  |
| CEE+MPA placebo | 196 | 11.31 ± 2.09 | (9.7, 11.3, 12.6) |  | 12.62 ± 5.48 | (8.9, 11.4, 15.5) |  |
| CEE+MPA | 203 | 11.03 ± 2.19 | (9.3, 10.7, 12.7) |  | 11.26 ± 4.89 | (8.1, 10.4, 14.1) |  |

### Total effects of air pollution exposures on MTL atrophy

Each interquartile range (IQR) increase in PM_2.5_ (IQR = 3.17 μg/m^3^) was associated with 0.29 (*95% CI= [−0.41, −0.18])* greater MTL atrophy, in standard deviation units. Increases in NO_2_ (per IQR 6.63 ppb) were also associated with greater atrophy in MTL volume, however, the magnitude of the effect was smaller (β*=-0.12; 95% CI= [−0.23, −0.02])*.

### AD-PS mediation models

Results of the SEMs examining the mediating roles of Alzheimer’s disease-related neurodegeneration are presented in Figure 3. Higher NO_2_ exposures were associated with larger AD-PS scores at MRI-1 (Figure 3 Panel A; β*=0.13; 95% CI= [0.03, 0.24]*). Accordingly, exposure-associated increase in AD-PS scores at MRI-1 partially mediated the effect of NO_2_ on MTL atrophy *(indirect effect* β*= −0.10; 95% CI= [−0.03, −0.01]),* explaining approximately 7% of the total effect of NO_2_. The AD-PS scores at MRI-1 did not significantly mediate observed associations between PM_2.5_ and MTL atrophy *(indirect effect* β*= −0.06; 95% CI= [−0.28, 0.01])*.

**Figure 3B** demonstrates mediation models with 5-year standardized change in AD-PS scores as the hypothesized mediator. Higher *PM_2.5_* exposures were associated with larger increases in AD-PS scores over time (β*=0.23; 95% CI= [0.12, 0.33]),* and larger *AD-PS score change* from MRI-1 to MRI-2 was associated with greater MTL atrophy (β*=−0.42; 95% CI=[−0.50, - 0.34]).* AD-PS change scores significantly mediated the effect of PM_2.5_ on MTL atrophy *(indirect effect* β*=−0.10; 95% CI= [−0.15, −0.05]),* explaining approximately 32% of the total effect. NO_2_ was not significantly associated with changes in AD-PS scores (β*=0.08; 95% CI=−0.03, 0.20]).* Changes in AD-PS score explained approximately 30% of the total effect of NO_2_ on MTL atrophy, although this estimated indirect effect was only marginally significant (β*= −0.04; 95% CI= [−0.09, 0.01])*.

**Figure 3.**
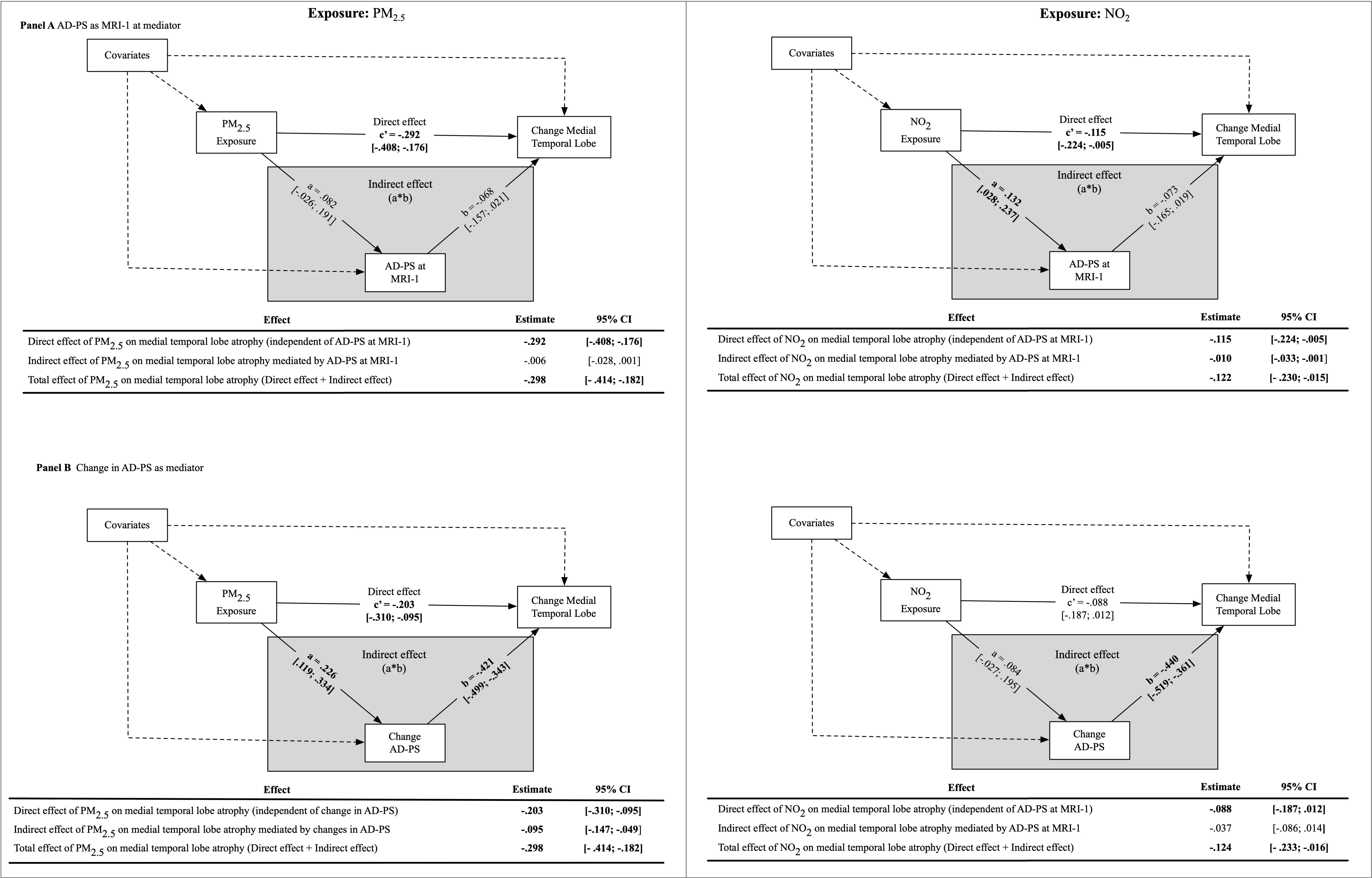
Results of mediation models examining whether Alzheimer’s disease related neurodegeneration (AD-PS) at Women’s Health Initiative Memory Study (WHIMS) MRI-1 (Panel A) or changes in AD-PS from MRI-1 to MRI-2 mediate associations between air pollution exposure on medial temporal lobe (MTL) atrophy (N = 627).

### WM-SVID mediation models

Higher NO_2_ was associated with lower WM-SVID volume at MRI-1 (β*=−0.10; 95% CI= [−0.18, −0.02])*, and was not associated with WM-SVID volume changes over the 5-year period. By contrast, PM_2.5_ was not significantly associated with WM-SVID volume at MRI-1, but higher PM_2.5_ was associated with less increases WM-SVID volume over time (β*=−0.20; 95% CI=[−0.30, −0.10])*. Neither WM-SVID volume at MRI-1, nor change in WM-SVID volume over time, mediated the effect of either exposure on MTL atrophy (Figure 4).

**Figure 4.**
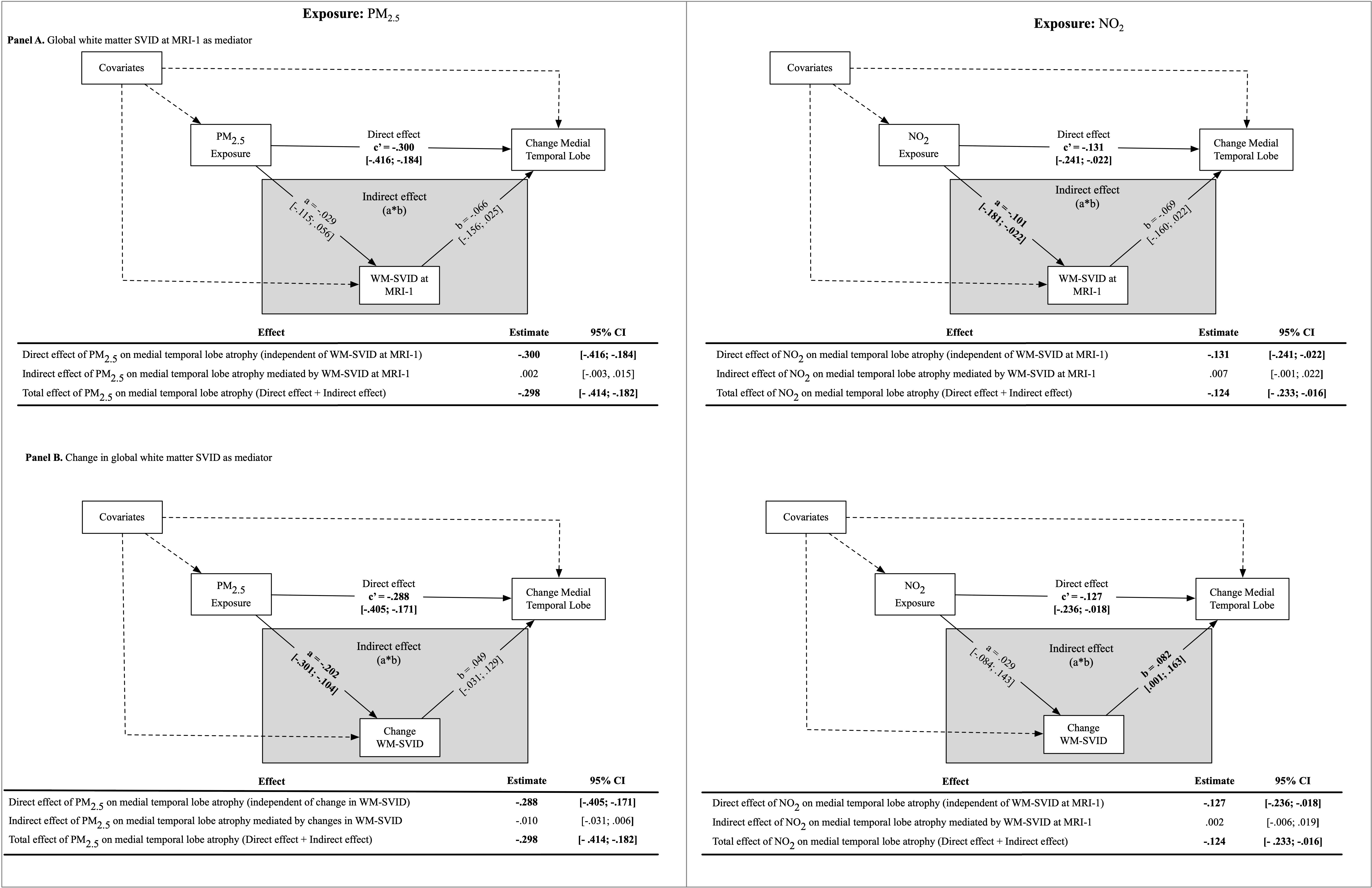
Results of mediation models examining whether global white matter small-vessel ischemic disease (WM-SVID) at Women’s Health Initiative Memory Study (WHIMS) MRI-1 (Panel A) or changes in global white matter small-vessel ischemic disease from MRI-1 to MRI-2 mediate associations between air pollution exposure on medial temporal lobe (MTL) atrophy (N = 627).

### Secondary analysis of MTL subregions

We observed a heterogeneous effect of air pollution exposures across MTL subregions. Higher exposures of both PM_2.5_ (β*=−0.44; 95% CI=[−0.56, −0.32])* and NO_2_ (β*=−0.20; 95% CI=[−0.32, −0.09])* were associated with greater PHG atrophy over the 5-year period, whereas greater ERC atrophy was only associated with higher exposure to PM_2.5_ (β*=−0.12; 95% CI=[−0.27, −0.01])*, but not NO_2_ (β*=−0.04; 95% CI=[−0.15, 0.06])*. Neither exposure was significantly associated with volume changes in the amygdala or hippocampus.

There was a significant indirect effect of NO_2_ on ERC atrophy that was mediated by AD-PS scores at MRI-1, explaining approximately 47% of the total effect (Supplemental Fig. 1A). Change in AD-PS partially mediated the total effect of PM_2.5_ on ERC atrophy, explaining over 63% of the total effect (Supplemental Fig. 1B). Changes in AD-PS scores explained approximately 15% of the total effect of PM_2.5_ on PHG atrophy (Supplemental Figure 2). AD-PS change scores did not mediate the effect of NO_2_ on the ERC or PHG.

## Discussion

The purpose of this study was to determine whether Alzheimer’s disease-related neurodegeneration or cerebrovascular damage explained the association between air pollution neurotoxicity and MTL atrophy in cognitively unimpaired older women. We found that higher NO_2_ and PM_2.5_ exposures were both associated with greater MTL atrophy over a 5-year period, which was partially explained by Alzheimer’s disease-related neurodegeneration but not WM-SVID. The effect of NO_2_ on MTL atrophy was only mediated by baseline AD-PS scores at MRI-1, whereas the effect of PM_2.5_ on MTL atrophy was only mediated by the change in AD-PS scores over time. Exploratory analyses showed that the mediation effect of AD-PS scores on exposure-related MTL atrophy was driven by cortical changes in the ERC and PHG, but not the amygdala or hippocampus. Collectively these results show that Alzheimer’s disease-related neurodegeneration, but not WM-SVID, contributes to MTL atrophy associated with late-life air pollution exposures in this sample of cognitively unimpaired older women.

Our results offer new insights into the explanatory mechanisms linking air pollution exposures to Alzheimer’s disease biomarkers and adverse cognitive outcomes that have been reported in prior work. The MTL is widely studied for its role in memory-related cognitive processes that can become impaired by Alzheimer’s disease neuropathological processes (e.g., neuroinflammation, lipid oxidation, abnormal beta-amyloid (Aβ) accumulation, tauopathy, neuronal damage, etc.). In rodents, air pollution exposures are associated with Alzheimer’s disease neuropathology in the MTL^43–50^, and contribute to impaired spatial learning and memory.^44,48,51–54^ Human studies also have linked exposures to Alzheimer’s disease biomarkers. Specifically, recent work from the Alzheimer’s & Families (ALFA) study revealed adverse associations. Higher NO_2_ and PM_2.5_ exposures were associated with higher brain fibrillar amyloid, as measured by higher centiloid values on positron emission tomography (PET) scans^55^ and with higher cerebrospinal fluid (CSF) levels of neurofilament light protein - a marker of neuronal injury. Ma et al.^56^ also linked PM_2.5_ exposures with CSF Aβ pathology in a large Chinese cohort of cognitively unimpaired adults.

Analyses of MTL subregions showed that the mediation effect of AD-PS change scores on PM_2.5_-related MTL atrophy was driven by cortical volume loss in the ERC and PHG, explaining over 60% and 15% of the total effects, respectively. This finding is consistent with numerous MRI studies that identify the ERC as one of the earliest brain regions to demonstrate structural damage and cell loss along the Alzheimer’s disease continuum.^57–59^ The ERC is also one of the earliest brain regions to exhibit pathological tau according to the original Braak & Braak model of Alzheimer’s disease neuropathology.^60^ Although the longitudinal effect of PM_2.5_ exposure on the ERC has not been studied in humans, mouse models of PM_2.5_ exposure have demonstrated increased levels of phosphorylated tau (p-tau) ^61^ and neuronal death^62^ in the ERC even at low levels of exposure.^50,61,63^ The PHG is also an initial site of preclinical brain atrophy and pathological tau deposition along the Alzheimer’s disease continuum^64^, yet our results showed that a large proportion (∼85%) of PHG atrophy associated with PM_2.5_ exposure could *not* be explained by Alzheimer’s disease-like neurodegeneration. The neuropathological processes and cellular mechanisms underlying these regional differences are unclear, although there is preliminary evidence suggesting the PHG may be more sensitive to aging and mixed pathologies that also impact the MTL^65^, such as primary age-related tauopathy (PART)^66^ and transactive response DNA-binding protein 43 (TDP-43).^67^ However, literature on the neuropathological distinctions between Alzheimer’s disease, PART, and TDP-43 is still emerging, and more work is needed to understand the role of air pollution exposures on brain atrophy and cognition in the context of mixed pathology neurodegeneration.

An interesting finding of this study was the unique mediating effects of *baseline* AD-PS scores on NO_2_-related MTL atrophy versus the unique mediating effects of AD-PS *change scores* on PM_2.5_-related MTL atrophy. These results partially align with prior work from our group showing that the adverse effects of PM_2.5_ on cognitive outcomes in older women were partially explained by AD-PS change scores only (not baseline AD-PS scores), though NO_2_ exposure effects were not examined.^23^ The underlying mechanisms that contribute to pollutant-specific effects are unknown, but could be due to differences in epigenetic changes that are initiated by NO_2_ versus PM_2.5_ exposure. For example, a large study of middle-aged Dutch individuals revealed significant associations between NO_2_ exposure and genome-wide DNA methylation at several CpG sites on genes that influence lung function, but there were no genome-wide DNA methylation associations with PM ^68^ In a separate epigenome-wide study of non-Hispanic White women in the U.S., higher NO_2_ exposure was associated with accelerated methylation-based “biological aging”, yet the effects of PM_2.5_ on biological aging depended on PM component profiles and the presence of other pollutants.^69^ Although the aforementioned studies did not focus on methylation patterns related to the brain, one possible interpretation of these studies and the results presented herein is that epigenetic changes activated by NO_2_ are involved in the initiation of Alzheimer’s disease pathological processes, whereas epigenetic and other neurobiological changes activated by PM_2.5_ are more closely linked to disease progression. In support of this hypothesis, the mediation effects of baseline AD-PS scores, rather than AD-PS change scores, on exposure-related MTL atrophy represent a stronger *causal* model of air pollution neurotoxicity since the predictor (exposures measured 3 years prior to MRI-1), mediator (baseline AD-PS at MRI-1) and outcome (MTL volume change over 5 years) variables were each measured at different time points.^43^ While differences in epigenetic changes offer an intriguing explanation for our pollutant-specific results, more work is needed to determine the role of pollutant-specific effects on DNA methylation in relation to brain integrity and Alzheimer’s disease-related neurodegeneration. Alternatively, NO_2_ and PM_2.5_ may simply represent surrogate markers for other pollutants with distinct neurotoxic properties.^70–73^

In contrast to the observed mediating effects of AD-PS scores on exposure-related MTL atrophy, WM-SVID was not significantly associated with and did not significantly mediate any of the observed associations between air pollution exposures and MTL atrophy in this study. Although the lack of association seems to contradict the extensive literature on the cardiovascular toxicity of airborne particles, several human studies have failed to find an association between exposures and MRI measures of WM-SVID.^16,19,74,75^ However, it is important to note that small vessel disease is an umbrella term for various pathologies of the brain microvascular system (e.g., lacunes, microbleeds, white matter hyperintensities (WMHs), enlarged perivascular spaces, etc.) that each requires unique neuroimaging techniques for detection.^76^ Prior studies (including the present study) of exposure-related SVD have focused on WMHs due to their high prevalence in older age and relative ease of measurement, but WMHs may not capture microvascular brain damage that results from air pollution neurotoxicity. The present study did not have access to alternative MRI markers of SVID that may mediate exposure-related MTL atrophy, but this is an important research direction for future work. The older age and generally good health of the sample may also indicate a survivor bias, such that individuals with significant WM-SVID would be deceased or cognitively impaired, and therefore would not meet the inclusion criteria for this study. We attempted to adjust for this potential bias in our analyses, but the lack of mediating effect remained.

This study has several major strengths. First, our longitudinal design allowed us to examine the mechanisms linking air pollution exposures to *intraindividual* changes in MTL volume, providing greater insight into the potential causal pathways of exposure-related MTL atrophy. Second, our richly phenotyped sample allowed us to adjust our mediation models for numerous potential confounding variables (e.g., socioeconomic, lifestyle, health factors, etc.) that have been associated with MTL atrophy in prior work. Third, the MUSE MRI protocol is another strength of our design as it uses an ensemble brain atlas for image segmentation that mitigates the impact of site-specific sources of noise. Finally, our serial mediation analyses allowed us to demonstrate the complete neuropathophysiological cascade linking exposures to increased, anatomical Alzheimer’s disease risk, greater MTL atrophy, and worse cognitive performance on tests of memory, which has not been done previously.

There are several limitations of this study that should also be acknowledged. First, results in this older all-female sample may not generalize to males or younger women. Participants were also in generally good health, which may limit generalizability to the broader population. Second, we cannot make inferences about the potential role of air pollution exposures on Alzheimer’s disease or SVID-related MTL atrophy during midlife as this study focused on neuroimaging associations with air pollution exposures during late life only. This may also explain the lack of a mediating effect of WM-SVID on MTL atrophy, as midlife vascular risk burden (rather than late life) is linked to greater late-life Alzheimer’s disease brain pathology^77^, hypoperfusion^78^, and brain volume.^79^ Third, our measure of anatomical Alzheimer’s disease risk did not account for individual differences in amyloid or tau positivity status as these biomarkers were not available for analysis. Fourth, we do not know whether structural changes in white matter contributed to exposure-related MTL atrophy as AD-PS scores were based on the spatial pattern of gray matter degeneration only. Finally, our measure of ambient air pollution exposures did not account for personal exposures at other locations (e.g., occupational), but it is unlikely this influenced the results as epidemiologic studies have shown that the associations between ambient exposure and health outcomes are not confounded by other sources of environmental pollutants.^80–83^

## Conclusions

Findings from the present study of cognitively unimpaired older women provide insight into the neuropathological processes by which ambient air pollution exposures contribute to MTL atrophy and subsequent cognitive difficulties. Our data support the role of AD-like neurodegeneration, but not SVID, as a putative pathway leading to exposure-related MTL atrophy over a 5-year period. The mediating effect of AD-like neurodegeneration varied across MTL subregions and explained the largest portion of the exposure effect on ERC atrophy, and only ∼30% of the total effect of air pollution exposure on total MTL atrophy, implying the importance of different neuropathologies. Future studies are needed to better understand the other neuropathological processes that explain associations between air pollution neurotoxicity and MTL atrophy in older adults.

## Data Availability

Data, codebook, and analytic code used in this report are held by the NIH-funded Coordinating Center of the Women's Health Initiative at the Fred Hutchinson Cancer Research Center and may be accessed as described on the Women's Health Initiative website: https://www.whi.org/page/working-with-whi-data.

## Abbreviations

Aβ: beta-amyloid
ADNI: Alzheimer’s disease neuroimaging initiative
AD-PS: AD pattern similarity
ADRD: Alzheimer’s disease and related dementias
ALFA: Alzheimer’s & Families
BMI: body mass index
CI: confidence interval
CVD: cardiovascular disease
ERC: entorhinal cortex
MCI: mild cognitive impairment
MTL: medial temporal lobe
MUSE: multi-atlas region segmentation
NO_2_: nitrogen dioxide
PART: primary age-related tauopathy
PHG: parahippocampal gyrus
PM_2.5_: particulate matter with aerodynamic diameter <2.5 μm
p-tau: phosphorylated tau
SEM: structural equation model
SES: socioeconomic status
TDP-43: DNA-binding protein 43
WHI: Women’s Health Initiative
WHIMS: Women’s Health Initiative Memory Study
WM-SVID: white matter small vessel ischemic disease

## Acknowledgements

We would like to acknowledge and thank Christos Davatzikos, Ph.D. and his research group at the University of Pennsylvania for providing the multi-atlas region segmentation (MUSE) processed estimates of the medial temporal lobe volume as well as the deep learning-based estimates of white matter small vessel ischemic disease.

## Supplementary material

Supplementary material is available at *Brain* online.

