## Supplementary figures and images for "Alzheimer’s Related Neurodegeneration Mediates Air Pollution Effects on Medial Temporal Lobe Atrophy"

### Supplemental

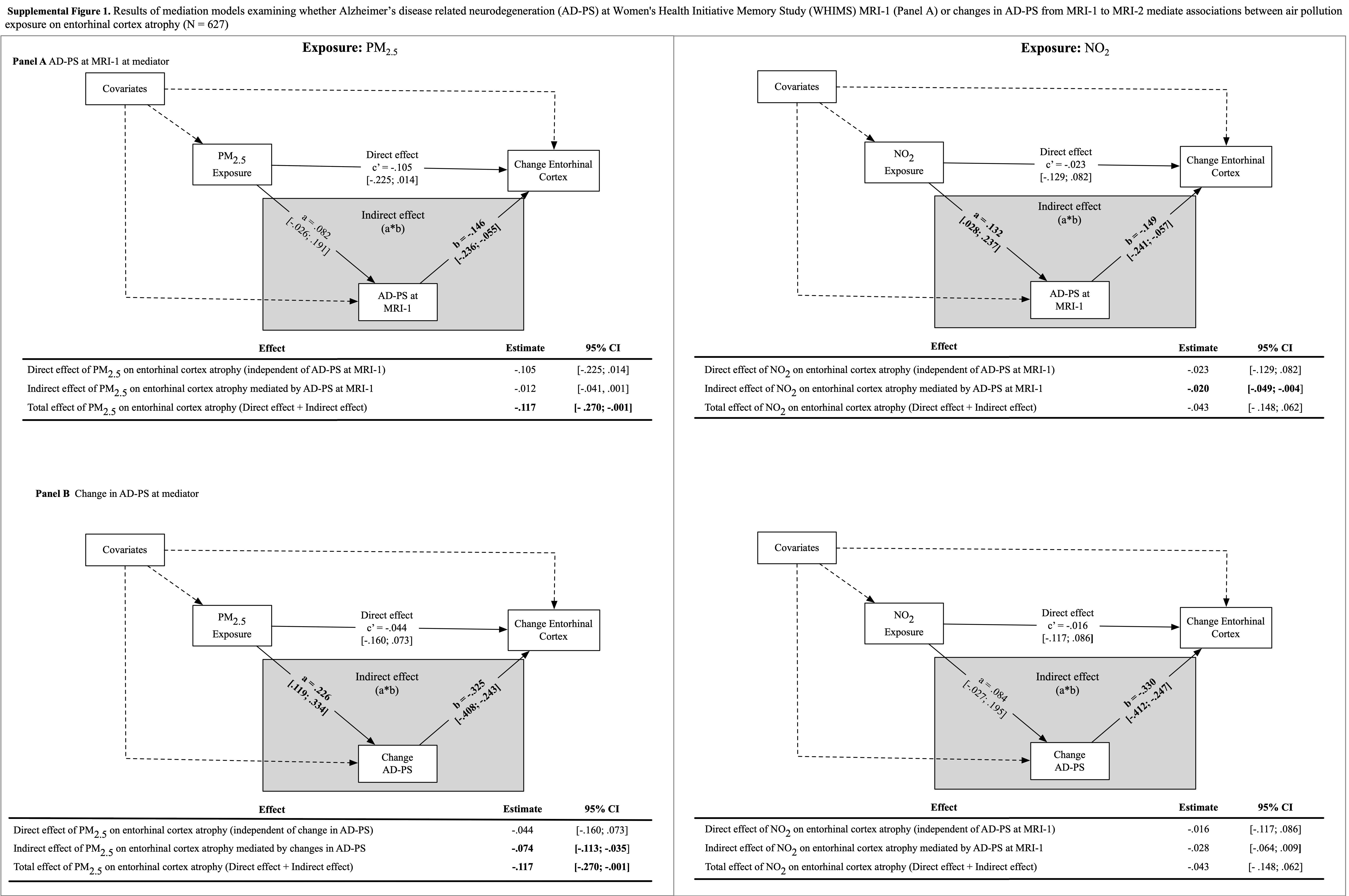
